# A Compressed Language Model Embedding Dataset of ICD 10 CM Descriptions

**DOI:** 10.1101/2023.04.24.23289046

**Authors:** Michael J. Kane, Casey King, Denise Esserman, Nancy K. Latham, Erich J. Greene, David A. Ganz

**Affiliations:** Department of Biostatistics, School of Public Health, Yale University, New Haven, USA; The Jackson School of Global Affairs Yale University, New Haven, USA; US Healthcare and Life Sciences Microsoft, Redmond, USA; Research Program in Men’s Health: Aging and Metabolism, Boston Claude D. Pepper Older Americans Independence Center for Function Promoting Therapies Brigham and Women’s Hospital, Boston, USA; Department of Medicine VA Greater Los Angeles/UCLA, Los Angeles, USA

**Keywords:** large language model, autoencoder, ICD-10-CM, electronic health records, EHR, NLP

## Abstract

This paper presents novel datasets providing numerical representations of ICD-10-CM codes by generating description embeddings using a large language model followed by a dimension reduction via autoencoder. The embeddings serve as informative input features for machine learning models by capturing relationships among categories and preserving inherent context information. The model generating the data was validated in two ways. First, the dimension reduction was validated using an autoencoder, and secondly, a supervised model was created to estimate the ICD-10-CM hierarchical categories. Results show that the dimension of the data can be reduced to as few as 10 dimensions while maintaining the ability to reproduce the original embeddings, with the fidelity decreasing as the reduced-dimension representation decreases. Multiple compression levels are provided, allowing users to choose as per their requirements. The readily available datasets of ICD-10-CM codes are anticipated to be highly valuable for researchers in biomedical informatics, enabling more advanced analyses in the field. This approach has the potential to significantly improve the utility of ICD-10-CM codes in the biomedical domain.

## Background

The International Classification of Diseases, 10th Revision, Clinical Modification (ICD-10-CM) [1] is a standardized classification system for categorizing diseases, disorders, and health conditions. ICD-10 was developed by the World Health Organization (WHO) and adapted for use in the United States as ICD-10-CM by the National Center for Health Statistics (NCHS) [2]. The standard plays a crucial role in the analysis of electronic medical records (EMRs) or electronic health records (EHRs) for several reasons:

1. Consistency and Standardization: The ICD-10-CM allows for a consistent and standardized method of coding and documenting medical conditions across healthcare providers and facilities. This helps to ensure accurate and uniform data exchange, analysis, and comparison.
2. Data Analysis and Research: The ICD-10-CM codes can be used to analyze patient data for clinical research, epidemiological studies, and public health surveillance. It helps to identify trends and patterns in diseases, monitor the effectiveness of treatments, and develop better prevention and management strategies.
3. Quality Measurement and Improvement: ICD-10-CM codes can be used to evaluate the quality of care provided by healthcare facilities, monitor patient outcomes and identify areas for improvement. This information can be used to enhance the overall healthcare delivery system.
4. Reimbursement and Billing: ICD-10-CM codes play a vital role in healthcare reimbursement by providing a standardized method to classify and report medical conditions. Insurance companies and other payers use these codes to determine appropriate payments for medical services rendered.
5. Health Policy and Planning: ICD-10-CM codes help health authorities and policymakers to identify population health needs, allocate resources, and develop targeted healthcare policies and interventions.

While ICD-10-CM codes do provide a consistent and comprehensive set of categories, their incorporation into statistical and machine learning analyses can be challenging for several reasons. First, in the 2019 version of the standard there are 71,932 categories with that number increasing in the 2022 version to 72,750 categories. As a result, analyses using these codes, where the set of codes is not restricted to smaller set, must take into account their high dimensionality or will require a large number of training samples in order to fit consistent models. Second, categorical variables are usually incorporated into analyses with a contrast encoding such as treatment, helmert, etc. Contrast numeric representations are orthogonal or, under appropriate statistical assumptions, independent with respect to their categories. However, ICD-10-CM codes represent a hierarchical structure, where codes are organized into chapters, blocks, and categories based on the type and anatomical location of the diseases or conditions. Applying traditional contrast encoding methods may not fully capture this hierarchical information, potentially resulting in a loss of valuable context and relationships between codes.

Researchers have considered alternative encoding methods or feature extraction techniques that can better represent the hierarchical structure of ICD-10-CM codes. However, incorporating both hierarchical structure and other contextual information in a general way can be difficult. The previous generation of word embeddings, which provide vector-encodings of words, were shown effective for these types of tasks, with models like `med2vec` [3] providing improved abilities to predict patient mortality; `inpatient2vec` [4] to predict clinical events; and `EHR2Vec` [5] to help analyze sequences of patient visits. Despite their advantages, word embeddings also have certain limitations. First, word embeddings are typically generated at the word or code level, and while word embeddings can capture semantic similarities, they ofen struggle to represent hierarchical representations like those found in ICD-10-CM codes. Second, traditional word embeddings generate a single vector for each word regardless of context. This means that the same code can have different meanings depending on where and when it is used. This is something these models do not capture. Third, word embeddings can have difficulty handling rare codes. Word embeddings typically require a sufficient number of training samples to learn meaningful representations. For rarely used ICD-10-CM codes, the learned embedding might not be reliable. Fourth, traditional word embedding provide static representations and do not change over time. However, in healthcare, the meaning and usages of certain codes can evolve, and these models cannot capture dynamic changes. Finally, the quality and representativeness of the word embeddings depend on the training data used to generate them. If the training data does not adequately cover the entire spectrum of medical conditions or encounters, the embeddings may not capture all relevant relationships or information.

Large language models (LLMs) address some of the shortcomings of traditional word embeddings through a combination of advanced techniques and architectures. Unlike traditional word embeddings that generate static representations, LLMs generate contextualized embeddings. These embeddings take into account the surrounding words or tokens, allowing for a more nuanced representation of words and codes in different contexts. This helps in capturing the semantic relationships between codes more effectively. These models are pre-trained on vast amounts of text data, allowing them to learn general language representations before being fine-tuned for specific tasks. This pre-training enables the models to leverage existing knowledge and adapt more effectively to new tasks, even with limited task-specific data. LLMs can be incrementally updated or fine-tuned with new data, allowing them to adapt to evolving medical knowledge and practices more effectively than static word embeddings. And, while not explicitly designed for hierarchical data like ICD-10-CM codes, LLMs can implicitly learn hierarchical relationships through their deep architectures and the context in which codes appear. This can help capture different levels of granularity and relationships between codes more effectively than traditional word embeddings.

Vector embeddings attempt to optimize the conditional probability of observing the actual output word given an input word (or vice versa, depending on the variant used). For instance, in the skip-gram variant, given a word *w*_*i*_ and a context word *w*_*j*_, the model is trained to maximize the following

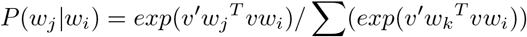

where *v*_*w*_ and 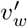 represent the “input” and “output” vector representations of a word w, and the summation in the denominator is over all words in the vocabulary. The vectors *v*_*w*_ and 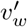 are the word embeddings learned by a similarity model.

LLM models also start by converting each word into an initial word embedding using an embedding matrix. However, these initial embeddings are then updated based on the context of the word. This is done by passing the embeddings through several layers of a transformer model, which uses self-attention mechanisms. The output of the transformer is a contextual embedding for each word. Mathematically, the self-attention mechanism can be represented as

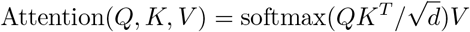

where *Q, K*, and *V* represent the query, key, and value matrices, which are derived from the input embeddings. The softmax function ensures that the weights of different words sum to 1, and the 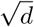 in the denominator is a scaling factor that improves the stability of the gradients during training. The resulting matrix product is a weighted sum of the value vectors, where the weights depend on the similarity between the query and key vectors.

To generate an embedding for a sentence or description, one common approach is to take the average of the contextual embeddings of the words in the sentence:

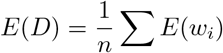

Here, *E*(*D*) is the embedding for the description, *E*(*w*_*i*_) is the contextual embedding for word *w*_*i*_, and the sum is over all words in the description.

The key difference between the two methods is that vector embeddings generate a single, static embedding for each word, while LLMs generates a dynamic, contextdependent embedding. This allows an LLM to capture nuances in meaning that can’t be represented with static embeddings.

There are several Bidirectional Encoder Representations from Transformers (BERT) or similar transformer-based models that have been used to generate embeddings for ICD-10-codes including ClinicalBERT [6, 7], BioBERT [8], and Med-BERT [9], but to our knowledge none of the current literature includes the applications of these models for the purpose of generating embeddings. That said, BERT has a maximum input length of 512 tokens. This would not be an issue for generating an embedding for a single ICD-10-CM code description. But for processing a long list of codes at once, or if one was to include additional context like patient notes or medical history, one could potentially exceed this limit.

LLM generated embeddings address many of these limitations. They take into account the surrounding words or tokens, allowing for a more nuanced representation of words and codes in different contexts. This helps in capturing the semantic relationships between codes more effectively. These models are pre-trained on vast amounts of text data, allowing them to learn general language representations before being fine-tuned for specific tasks. This pre-training enables the models to leverage existing knowledge and adapt more effectively to new tasks, even with limited task-specific data. LLMs can be incrementally updated or fine-tuned with new data, allowing them to adapt to evolving medical knowledge and practices more effectively than static word embeddings. And, while not explicitly designed for hierarchical data like ICD-10-CM codes, LLMs can implicitly learn hierarchical relationships through their deep architectures and the context in which codes appear. This can help capture different levels of granularity and relationships between codes more effectively than traditional word embeddings.

This paper describes data sets provided as `.csv` files, which are available online, ICD-10-CM codes to embeddings (a numeric vector of values), based on their descriptions. The embeddings were generated using the BioGPT LLM [10],which was trained on the biomedical literature including PubMed [11], PubMed Central [12], and clinical notes from MIMIC-III [13]. This model was shown to be superior at encoding context and relational information than competitors in the medical domain. Since the dimension of the embedding LLM is relatively high (42,384), we provide dimension-reduced versions in 1,000, 100, 50, and 10 dimensions. The model generating the data were validated in two ways. The first way validates the dimension reduction. The embedding data were compressed using an auto-encoder. The out-of-sample accuracy of a validation set is examined as well as the performance of the model for other versions (by year) of the ICD-10-CM specification. Our results show that we can reduce the dimension of the data down to as few as 10 dimensions while maintaining the ability to reproduce the original embeddings, with the fidelity decreasing as the reduced-dimension representation decreases. The second way validates the conceptual representation by creating a supervised model to estimate the ICD-10-CM hierarchical categories. Again, we see as the dimension of the compressed representation decreases the model accuracy decreases. Since multiple compression levels are provided, users are free to choose whichever suits their needs, allowing them to trade off accuracy for dimensionality.

The paper proceeds as follows. The next section provides a high-level description of the BioGPT and the embedding along with the construction of the autoencoder used to reduce the dimension of the embedding representation. That section then provides validation for both the dimension reduction as well as the representation. The third section provides an example of how to use the dataset to cluster ICD-10-CM codes using the R programming environment [14]. The final section provides a broader look at the incorporation of LLM approaches to these types of data.

The data sets and code to generate them are available in a public repository on Github ^[1]^. The data are licensed under the Creative Commons Attribution NonCommercial ShareAlike 4.0 International License ^[2]^. The code is licensed under GPL-v2 ^[3]^.

## Construction and content

The provided data are generated by embedding ICD-10-CM descriptions using the BioGPT-Large model, which comprises 1.5 billion parameters and is accessible via the Hugging Face model repository, ^[4]^ and then performing a dimension reduction using an autoencoder. The embedding process involves tokenizing textual phrases into tokens (words, subwords, or characters) and mapping them to unique vocabulary IDs. Token IDs are passed through an embedding layer, resulting in a sequence of continuous embedding vectors. Positional encodings are added elementwise to these vectors, enabling the model to capture token order and relative positions. The embeddings are then contextualized by passing them through the model’s layers. An attention mask selectively controls information flow in the attention mechanism, allowing the model to weigh the importance of input tokens when generating contextualized embeddings in a 42,384-dimension space.

The embedding is then compressed using an autoencoder. The autoencoder used here is a series of fully connected layers where the number of hidden nodes is approximately one order of magnitude smaller than the previous layer and then an order of magnitude larger until the output layer. For example, the autoencoder compressing to 10 dimensions has layers of size 42,384, 1,000, 100, 50, 10, 50, 100, 1,000, 42,384. Models whose dimension is large use the same structure while retaining only the appropriate layers.

### Validating the dimension reduction

The autoencoder compressing the LLM embedding was fit on the 2019 ICD-10-CM descriptions for 20 epochs, with batch sizes 64, 128, and 256, mean-square error loss between the embedding and autoencoder estimate, and a validation data set comprised of random subset of 10% of the samples. The model performance is shown in Table 1. Based on these results the models with the best validation loss for each of the compressed embedding dimensions selected for further validation and eventual distribution.

**Table 1.** The autoencoder parameters and performance ordered by increasing validation loss.

| Embedding Dimension | Batch Size | Training Loss | Validation Loss |
| --- | --- | --- | --- |
| 100 | 64 | 0.534 | 0.339 |
| 100 | 128 | 0.487 | 0.381 |
| 50 | 256 | 0.403 | 0.392 |
| 1000 | 64 | 0.542 | 0.402 |
| 100 | 256 | 0.556 | 0.444 |
| 1000 | 128 | 1.073 | 0.486 |
| 10 | 256 | 0.599 | 0.594 |
| 10 | 128 | 0.628 | 0.609 |
| 10 | 64 | 0.679 | 0.641 |
| 50 | 64 | 1.134 | 0.699 |
| 1000 | 256 | 30.435 | 0.803 |
| 50 | 128 | 1.053 | 0.894 |

In addition to the 2019 validation the models selected for distribution were tested on the 2020-2022 data sets to ensure their performance is comparable over years. The results are shown in Table 2. It should be noted that the ICD-10-CM codes do not vary much from one year to the next, so we should not expect large differences. As expected, the mean square error and coefficients of determination are similar to the 2019 data.

**Table 2.**
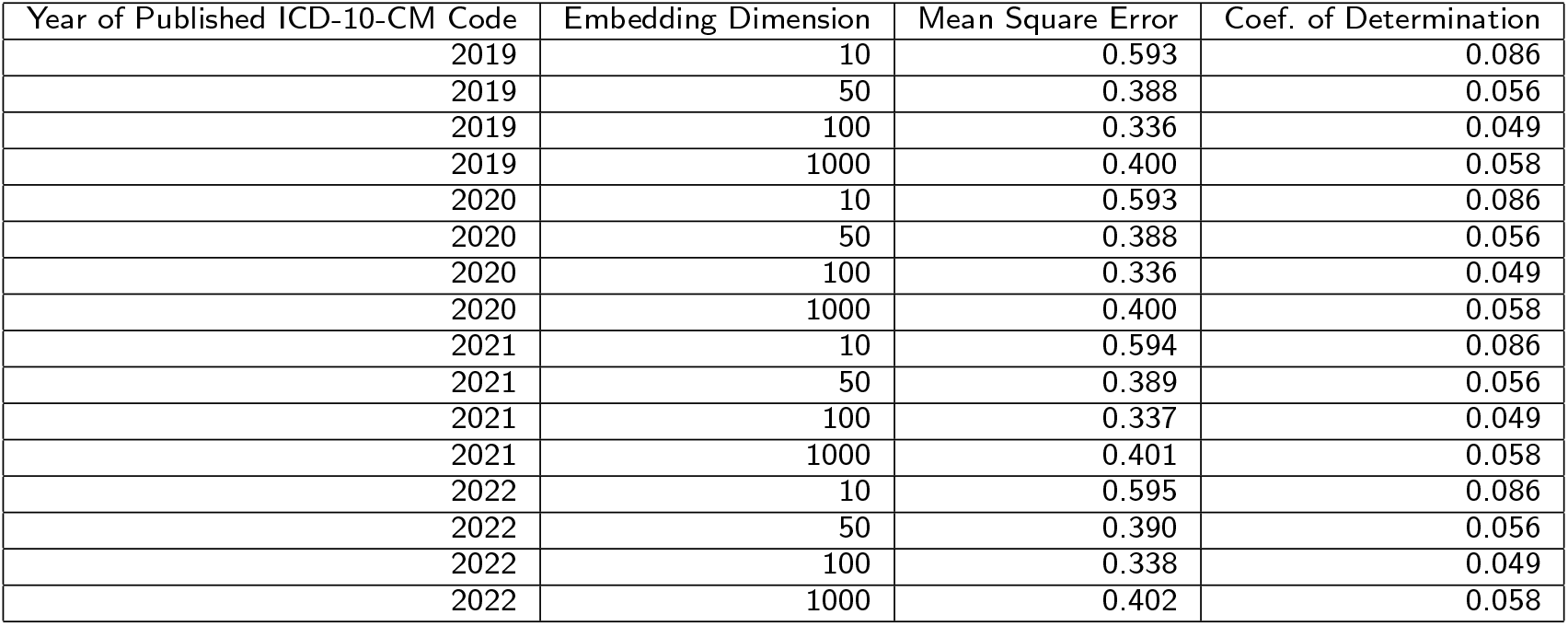
The autoencoder validation performance ordered by year.

| Year of Published ICD-10-CM Code | Embedding Dimension | Mean Square Error | Coef. of Determination |
| --- | --- | --- | --- |
| 2019 | 10 | 0.593 | 0.086 |
| 2019 | 50 | 0.388 | 0.056 |
| 2019 | 100 | 0.336 | 0.049 |
| 2019 | 1000 | 0.400 | 0.058 |
| 2020 | 10 | 0.593 | 0.086 |
| 2020 | 50 | 0.388 | 0.056 |
| 2020 | 100 | 0.336 | 0.049 |
| 2020 | 1000 | 0.400 | 0.058 |
| 2021 | 10 | 0.594 | 0.086 |
| 2021 | 50 | 0.389 | 0.056 |
| 2021 | 100 | 0.337 | 0.049 |
| 2021 | 1000 | 0.401 | 0.058 |
| 2022 | 10 | 0.595 | 0.086 |
| 2022 | 50 | 0.390 | 0.056 |
| 2022 | 100 | 0.338 | 0.049 |
| 2022 | 1000 | 0.402 | 0.058 |

### Validating the embedding representation

As a final step in the validation process, we use the fact that in addition to the description, the ICD-10-CM codes themselves carry hierarchical information, which can be used to ensure that conceptual relationships are preserved in the compressed embeddings. In particular, the leading letter and two numeric values categorize codes. For example, codes A00-B99 correspond to infectious and parasitic diseases, C00-D49 correspond to neoplasms, etc. We can therefore ensure that at least some of the relevant relationships are preserved in the compressed embedding representation by confirming that the categories can be estimated at a rate higher than chance using a supervised model. Furthermore, we can quantify how much relevant predictive information is lost in lower-dimensional representations.

The training data consists of a one-hot encoding of the ICD-10-CM categories as the dependent variable and the compressed embedding values as the values. The model consists of two hidden layers with 100 nodes each. The loss function selected was categorical cross-entropy. The model was trained using 30 epochs and a validation data set comprised of 10% of samples, chosen at random. The performance in terms of both the accuracy and the balanced accuracy is shown in Table 3. As with most problems of this type, compression of the data corresponds to a decrease predictive information.

**Table 3.** The supervised models’ performance ordered by increasing embedding dimension.

| Embedding Dimension | Accuracy | Balanced Accuracy |
| --- | --- | --- |
| 10 | 0.815 | 0.698 |
| 50 | 0.925 | 0.873 |
| 100 | 0.935 | 0.891 |
| 1000 | 0.960 | 0.927 |

Of note, the goal in presenting these results is not to necessarily to maximize the prediction accuracy. Rather, it is to show that the embedding retains the hierarchical information in the ICD-10-CM codes. Some of the codes correspond to conditions that could be classified in several ways, and as a result coding for at least some of the conditions might be considered non-systematic. Based on this criterion, we can conclude the embedding does retain much of the structural and conceptual information denoted in the descriptions, at least in terms of mapping to key categories of diseases and conditions.

## Utility and discussion

To illustrate the utility of the data, we present a simple example of how one might use the embedding information in the R programming environment and making use of the `dplyr` [15], `ggplot2` [16], `readr` [17], `Rtsne` [18], and `stringr` [19] packages. Suppose we would like to visualize the ICD-10-CM codes beginning with G (diseases of the nervous system), I (diseases of the circulatory system), J (diseases of the respiratory system), and K (diseases of the digestive system) to better understand the contextual relationships between these categories or specific conditions in the the 50-dimensional embedding. For convenience, the projects page includes an `.rds` file containing the available embeddings along with their URLs, which can be retrieved from the R console. The code categories can then be visualized by performing another dimension reduction (in this case we will use the `Rtsne` package), to 2 dimensions that can be presented as a scatter plot.

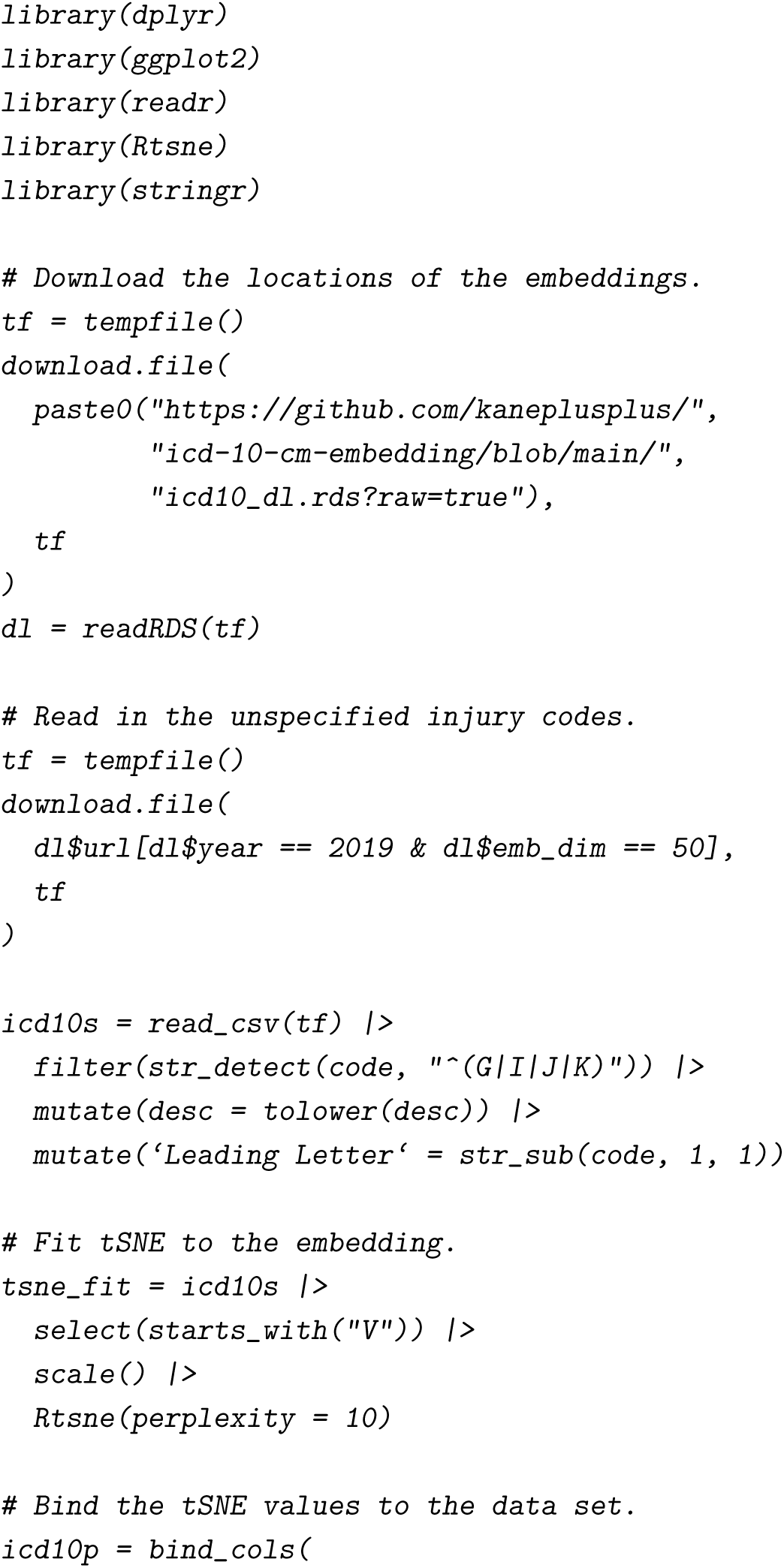
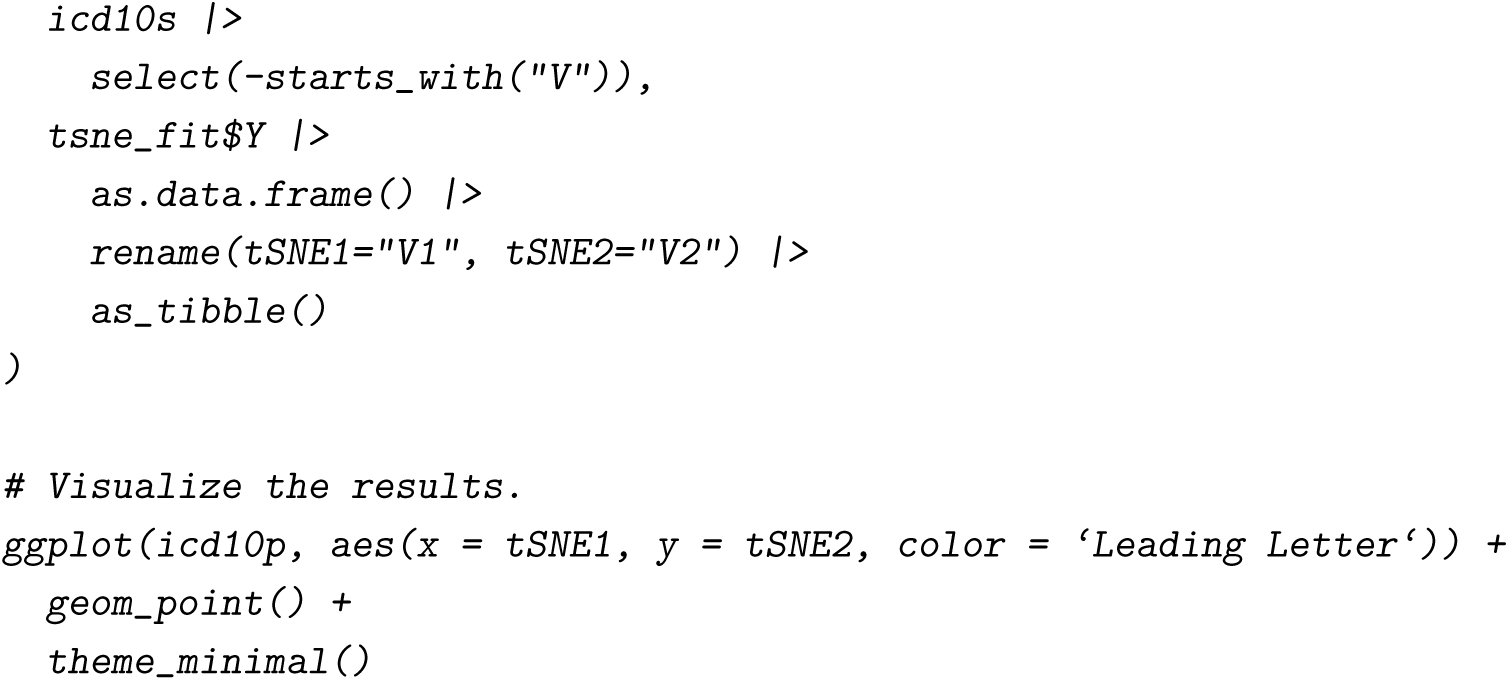

The output visualization is presented in Figure 1 and shows that a subset of the circulatory diseases (I) and nervous system diseases (G) are well-differentiated from other conditions. It also shows overlap between other conditions related to K (digestive diseases), J (respiratory diseases), and I (circulatory). While this approach is effective, there are some challenges of which we should be aware. While not insurmountable, they are as follows:

1. Interpretability: A significant challenge in machine learning, particularly with complex models like large language models and autoencoders, is interpretability. In healthcare, the ability to understand and explain why a model makes a particular prediction is crucial. This could impact patient trust, clinician adoption, and even legal and regulatory compliance. Techniques like LIME (Local Interpretable Model-Agnostic Explanations) or SHAP (SHapley Additive exPlanations) can be used to improve interpretability, but they don’t provide perfect solutions and can be computationally expensive.
2. Overfitting: Overfitting is a common issue in machine learning where a model learns the training data too well and performs poorly on unseen data. This can be particularly problematic in healthcare, where the stakes are high. Techniques such as cross-validation, regularization, or dropout layers can be used to prevent overfitting.
3. Data Privacy: Patient data is highly sensitive, and its usage is strictly regulated (e.g., by laws like HIPAA in the US). Even if the data used to generate the embeddings is anonymized, the model must be carefully designed and used to avoid potential privacy leaks.
4. Generalizability: A model trained on one dataset may not perform well on another due to differences in population characteristics, data collection methods, etc. Ensuring that models generalize well across different settings is a significant challenge.
5. Quality of Input Data: The quality of the embeddings depends heavily on the quality of the input data. If the descriptions associated with the ICD-10-CM codes are inaccurate or not comprehensive, the resulting embeddings may also be flawed. This is a fundamental issue in any data-driven approach: “garbage in, garbage out.”
6. Capturing Hierarchical Structure: The ICD-10-CM coding system has a hierarchical structure where certain codes are nested within broader categories. While embeddings generated from code descriptions may capture semantic meaning, they might not inherently capture this hierarchical structure. Advanced techniques or additional processing might be required to represent this structure effectively. This challenge is the one most specific to ICD 10 codes. An LLM trained to generate embeddings from descriptions can capture semantic similarity between different descriptions, but it might not explicitly capture the hierarchical structure. For example, I might recognize that “hypertension” and “cardiovascular disease” are related concepts, but it might not understand that hypertension is a specific type of cardiovascular disease.

**Figure 1.**
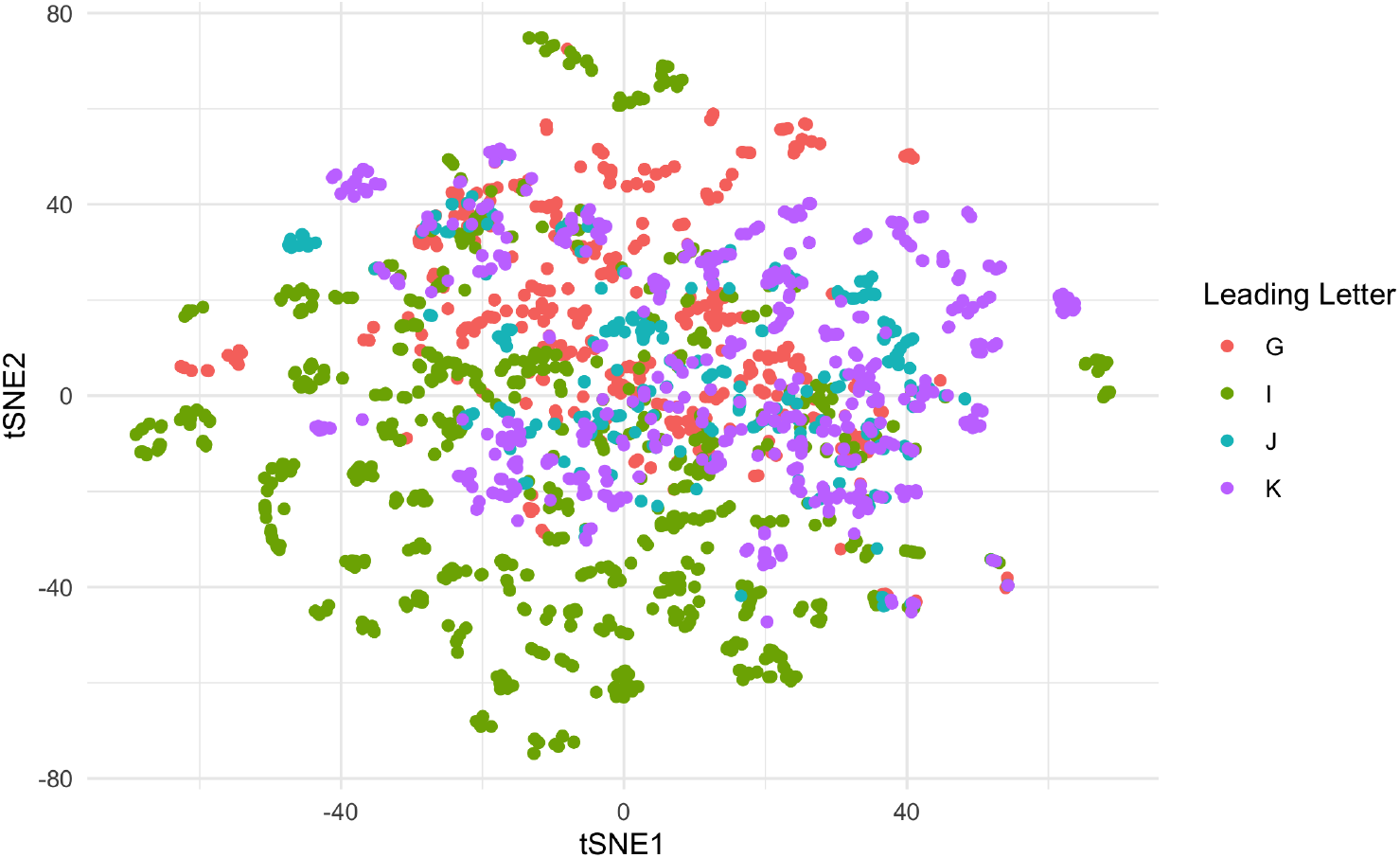
The tSNE plot of the codes.

## Conclusions

This paper presents novel datasets offering numerical representations of ICD-10-CM codes by generating description embeddings using a large language model and applying autoencoders for dimensionality reduction. The approach is versatile, capable of handling categorical variables with numerous categories across various domains. By capturing relationships among categories and preserving inherent information, the embeddings serve as informative input features for machine learning models. The readily available datasets are anticipated to be highly valuable for researchers incorporating ICD-10-CM codes into their analyses, retaining contextual information. This approach has the potential to significantly improve the utility of ICD-10-CM codes in biomedical informatics and enable more advanced analyses in the field. Data analysts can easily incorporate them into their own analyses by substituting the embedding values for other, lower-information representations including the categorical ones described above to derive the benefits of the conceptual information encoded in their embedding. Future work will address some of the challenges of capturing hierarchical structure in ICD-10-CM coding systems, experimenting with Ontology-based methods, hierarchical clustering, hierarchial autoencoding, graph neural networks and incorporating hierarchical information in training.

## Data Availability

All data produced are available online at https://github.com/kaneplusplus/icd-10-embedding.

## Competing interests

The authors declare that they have no competing interests.

### Ethics approval

Not applicable.

### Consent to participate

Not applicable.

### Consent for publication

Not applicable.

### Availability of data and materials

All data presented here along with documentation for reproducing presented materials is available at https://github.com/kaneplusplus/icd-10-cm-embedding.

### Code availability

All code presented here along with documentation for reproducing presented materials is available at https://github.com/kaneplusplus/icd-10-cm-embedding.

### Authors’ contributions

Kane proposed, implemented, and created the dataset and wrote the article. Ganz provided direction for the research and validated results manually. King provided assessment of the model, a detailed analysis of the limitations of vector based and BERT approaches, a discussion of LLM limitations and feedback. Esserman, Latham, and Green provided feedback and made suggestions through the entire process.

## 0.1 Acknowledgements

Not applicable.

## Footnotes

[1] https://github.com/kaneplusplus/icd-10-cm-embedding

[2] https://creativecommons.org/licenses/by-nc-sa/4.0/legalcode

[3] https://www.gnu.org/licenses/old-licenses/gpl-2.0.en.html

[4] https://hugginface.co

